# The socioeconomic patterning of Great-Britain’s retail food environment: A repeated cross-sectional study of area-level deprivation and food outlet density from 2011-2024

**DOI:** 10.1101/2025.03.07.25323573

**Authors:** Alexandra Boskovic, Thomas Burgoine, Jody Chantal Hoenink

**Author notes:** Corresponding author: Dr Thomas Burgoine, MRC Epidemiology Unit, University of Cambridge School of Clinical Medicine Box 285 Institute of Metabolic Science, Cambridge Biomedical Campus, Cambridge, CB2 0QQ, United Kingdom.

## Abstract

**Background:** Although previous research has demonstrated socioeconomic differences in the distribution of food outlets, no study to date has examined these patterns across the entire territory of Great Britain (GB) over time. This study provides an up-to-date repeated cross-sectional analysis into changes in GB’s retail food environment from 2011 to 2024 and its socioeconomic distribution.

**Methods:** Coordinates of all fast-food outlets and supermarkets in GB from the Ordnance Survey Points-of-Interest data were used to calculate outlet densities within small areas, adjusted by yearly population estimates and linked to Index of Multiple Deprivation measures. Counts of the different food outlets were combined in the modified Retail Food Environment Index (mRFEI). Multi-level linear regression and beta-regression models were used to assess associations between deprivation and the retail food environment (fast-food outlets, supermarkets and mRFEI).

**Results:** In GB, fast-food outlet density increased by 36%, supermarket density increased by 17% and mRFEI decreased by 5% from 2011 to 2024. More deprived areas were associated with greater fast-food outlet density and lower mRFEI in all years compared to less deprived areas; the gap in fast-food outlet density between the most and least deprived areas widened by 28% from 2011-2024. There were no statistically significant differences in supermarket density by area-level deprivation.

**Conclusions:** Our findings indicate that higher densities of fast-food outlets in more deprived areas are not offset by equivalent access to supermarkets offering healthier foods, potentially limiting healthier dietary choices for residents in these communities. Given the links between food outlet exposure and dietary behaviours in GB, policies aiming to improve dietary outcomes should prioritize more deprived areas, thereby contributing to the reduction of socioeconomic inequalities in the retail food environment, diet, and health outcomes.

## Introduction

The physical food environment may play an important role in shaping dietary behaviours and health [1,2]. Neighbourhood retail food environments (RFEs), which include supermarkets, convenience stores and fast-food outlets, contribute to the landscape of available food options. When these environments are characterized by high densities of unhealthy food options and limited availability of healthier choices, they can foster dietary patterns associated with excess energy consumption and obesity [3–5].

Multiple studies from the United Kingdom (UK) found a greater density of fast-food outlets in more deprived compared to less deprived areas [6–8]. This pattern may partially explain the higher obesity rates observed in socioeconomically disadvantaged communities, given evidence linking fast-food availability to consumption patterns and obesity risk [9–11]. However, the literature on supermarkets—their distribution and accessibility across the socioeconomic spectrum—is less consistent; some studies find greater supermarket access in more deprived areas [12], others find less [13] and others report no difference [8]. These mixed findings underscore the complexity of understanding how different components of the RFE may contribute to diet-related health disparities. Most existing studies have used cross-sectional data, limiting insights into both how RFEs have changed over time and their potential role in widening health inequalities. One repeated cross-sectional study in Norfolk, England, found that the most deprived areas experienced a 43% increase in fast-food outlets from 1990-2008, while the least deprived areas saw a 30% increase. Supermarkets experienced a 29% increase and although more deprived areas had higher supermarket density over the study period, there were no significant differences by area deprivation [8]. However, this study was constrained to a geographically limited study area with potential implications for generalisability to other regions of the UK and is relatively out-of-date. The authors also examined different aspects of food retail in isolation, as opposed to considering a combined measure of the RFE [8].

Increasingly, researchers are using both relative and absolute RFE measures to assess the impact of food environments on health [14,15]. For example, the modified Retail Food Environment Index (mRFEI) presents healthy food outlets as a proportion of all food outlets, and may help determine whether the presence of healthy food options could offset exposure to unhealthy outlets [15,16]. However, temporal analysis of relative RFE measures at the area-level in relation to deprivation remains largely unexplored.

In this study, we addressed these knowledge gaps by conducting an up-to-date, nationwide analysis of temporal trends in the socioeconomic patterning of RFEs. We analysed changes in supermarket and fast-food outlet density at the small-area level from 2011 to 2024 using absolute and relative measures across GB (England, Scotland and Wales) and assessed RFE changes by deprivation.

## Methods

In this repeated cross-sectional study, Ordnance Survey Points-of-Interest (OS PoI) data were used to source locations of all supermarkets and fast-food outlets in GB in each year 2011-2024. Counts of outlets were generated and converted to densities using population estimates. The proportion of supermarkets compared to the total retail food environment was calculated using the mRFEI. Associations between area-level deprivation, year, food outlet density and mRFEI were examined.

### Food outlet data

The addresses, coordinates and names of all fast-food outlets and supermarkets in GB, from 2011-2024, in September for 2011-2013, December for 2014-2023 and June for 2024, were sourced from OS PoI [17,18]. All outlets in the classes “Fast Food and Takeaway Outlets”, “Fast Food Delivery Services” and “Fish and Chip Shops” were included as fast-food outlets. All outlets in the class “Supermarket Chains” were included as supermarkets, in addition to outlets selected by name from the “Convenience Stores and Independent Supermarkets” and “Frozen Foods” classes, based on market share, total till sales and shopper’s preferred store [19–22] Supermarkets included in this study were Sainsbury’s, Tesco, The Co-operative, Morrisons, Iceland, Marks & Spencer Simply Food, Booths, Waitrose, TFC, Asda, CK’s Supermarkets, Aldi, Heron Foods and Filco. OS PoI data contained duplicates, likely due to food outlets classified in multiple categories. Thus, duplicates were removed by first screening outlets with identical names and geographical locations. Then, duplicates with the same location by postcode but slight variations in names, e.g. Tesco and Tesco Express, were removed.

### Middle Super Output Area and Intermediate Zone boundaries

Small areas were defined as the 2011 Middle Super Output Area (MSOA) for England and Wales and the Intermediate Zone (IZ) for Scotland. MSOAs and IZs include 5000-15,000 residents. The study area (GB) comprised 6856 MSOAs in England, 408 in Wales, and 1279 IZs in Scotland. Area boundaries were sourced from the Office for National Statistics (ONS) and the Scottish government’s spatial data website [23,24].

### Deprivation data

The English, Scottish and Welsh Indices of Multiple Deprivation (IMD), henceforth referred to collectively as IMD, were used as measures of area-level deprivation. IMD is a relative measure of area deprivation, which although calculated differently for each nation, is based on shared weighted factors such as income, education, and crime [25–27]. IMD data are not native to MSOAs or IZs, and therefore IMD was calculated as the mean of scores across smaller Lower Super Output Areas (England and Wales) and Data Zones (Scotland) contained within them. IMD scores were then ranked and divided into country-specific quintiles. IMD data are updated every 3-4 years, and so we used IMD data from multiple years across our study period (2011-2024). For England, 2015 IMD was used for years 2011-2018, and 2019 data for years 2019-2024. For Scotland, 2016 IMD was used for years 2011-2019, and 2020 for years 2020-2024. For Wales, 2014 IMD was used for years 2014-2018, and 2019 data for years 2019-2024.

### Covariates

Covariates were small area urban-rural classification and age– and gender-stratified population. Urbanicity has previously been associated with higher food outlet density in a UK study, and was therefore included as a covariate [12]. Additionally, the proportion of the population who were male, and the population under 30 were included due to previous findings of higher fast-food consumption in these groups [5,28]. Resident population data and area urban/rural classification were available from ONS (England and Wales) and the Scottish Government website [29,30]. Urban-rural classification was aggregated from smaller LSOA/DZ boundaries based on the modal classification, and with bimodal MSOAs/IZs classified as urban.

### Outcome Measures

Annual counts of fast-food outlets and supermarkets within MSOAs/IZs from 2011-2024 were generated in ArcGIS Pro [31] and transformed into densities per 10,000 population per MSOA/IZ. Outliers (mean + 3* standard deviation) were removed, mainly city-centres of large GB cities. Our relative measure of RFE healthfulness, mRFEI, was calculated as the total number of supermarkets divided by the total number of all food outlets, multiplied by 100. Areas with no food outlets were removed prior to analysis of mRFEI (6.8% removed). The final sample size consisted of 8417 areas for food-outlet outcome analysis and 8158 areas for mRFEI outcome analysis.

### Statistical Analysis

Descriptive statistics included the mean fast-food outlet or supermarket count per 10,000 population and mRFEI per small area, by quintile of deprivation from 2011-2024. We employed linear multi-level regression models to examine associations between deprivation quintile and food outlet counts (per 10,000 population per MSOA/IZ) including interaction terms between year and deprivation quintile. These models accounted for repeated measures over time clustered within MSOAs/IZs and were adjusted by covariates. For the mRFEI outcome, we employed a beta regression model instead as the outcome was bounded between 0 and 1. Coefficients were transformed into predicted outcomes with 95% confidence intervals (CI) using estimated marginal means. Analyses were conducted in R version 4.4.0 [32].

## Results

### Descriptive statistics

Between 2011 and 2024, the mean fast-food outlet density in GB rose by 35.7%, from 7.0 to 9.5 fast-food outlets per 10,000 population (Figure 1). Since 2014, fast-food outlet density has consistently increased, with the largest yearly increase occurring from 2020 to 2021, at a mean of 0.4 fast-food outlets per 10,000 population. Supermarket density also increased over the 14-year period, rising by 16.7%, from 1.8 in 2011 to 2.1 in 2024. On average, all years, except for 2015-2016 and 2018-2019, saw increases in supermarket density, with the largest yearly increase occurring from 2021 to 2022, at 0.1 supermarkets per 10,000 population. The mean mRFEI per MSOA/IZ decreased by 5.4%, from 23.9% in 2011 to 22.6% in 2024, showing that supermarkets were becoming a smaller proportion of all food outlets (Supplementary Figure 1).

**Figure 1.**
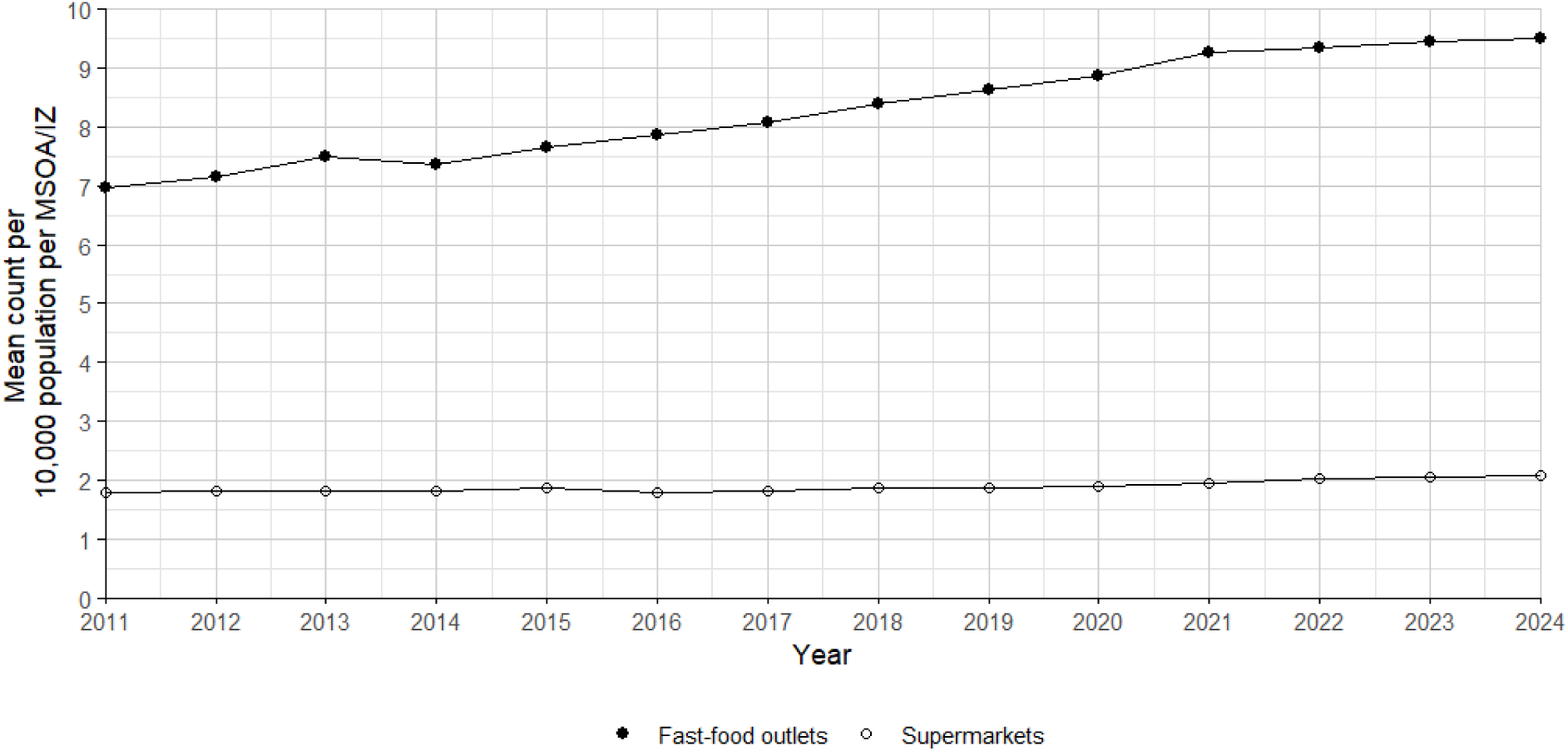
Mean fast food outlet and supermarket count per 10,000 population per MSOA/IZ from 2011-2024 in GB.

### Fast-food density by area-level deprivation

Overall, fast-food outlet density increased over the study period across all levels of deprivation (Figure 2). The most deprived areas (Q1) had the greatest density of fast-food outlets at each time point, and the least deprived areas (Q5) had the smallest density of fast-food outlets at each time point. These differences were statistically significant as indicated by the non-overlapping 95% CI. From 2011 to 2024, the most deprived areas (Q1) experienced an increase in number of fast-food outlets per 10,000 population of 3.6, rising from 9.9 to 13.5, while the least deprived MSOAs/IZs experienced the smallest increase at 1.9, rising from 3.9 to 5.8 outlets per 10,000 population (Supplementary Table 1). The gap in fast-food outlet density between the least and most deprived quintiles widened by 28.3%, from 6.0 in 2011 to 7.7 in 2024 (Supplementary Table 1).

**Figure 2.**
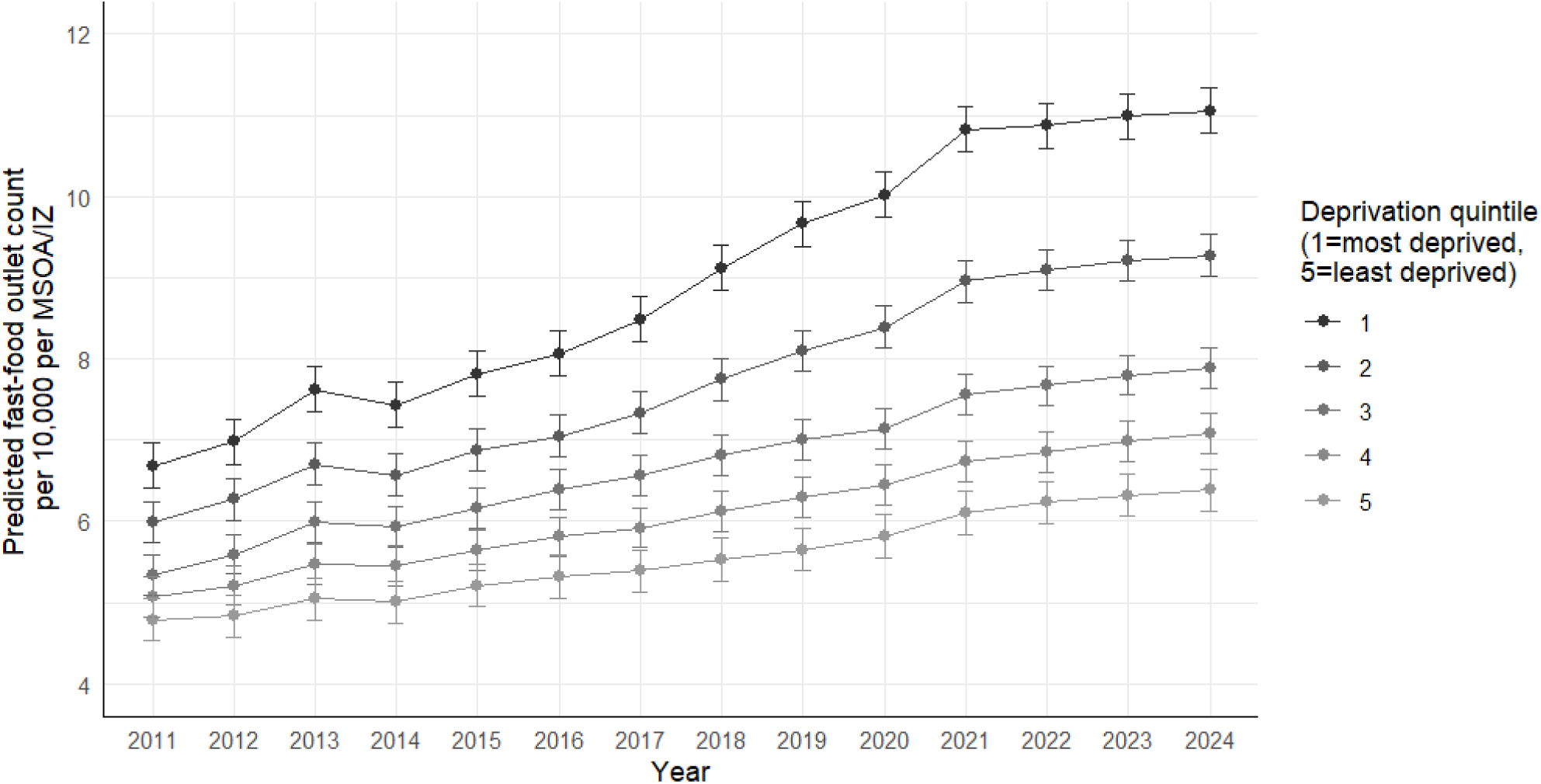
Mean predicted fast-food outlet counts per 10,000 population by deprivation quintile in GB (2011–2024) with 95% CI. Coefficients were derived from linear multi-level models with interaction terms between year and deprivation quintile. Predicted fast-food densities were adjusted for the effect of proportion of population under age 30, proportion of population male and urban/rural classification.

### Supermarket density by area-level deprivation

When stratified by area-level deprivation, the 95% CI for supermarket density estimates largely overlapped across deprivation quintiles, suggesting that differences between these groups were not statistically significant (Figure 3). While there was a general upward trend in supermarket density, this change was not uniform over time or consistently distinct across levels of deprivation (Supplementary Table 1).

**Figure 3:**
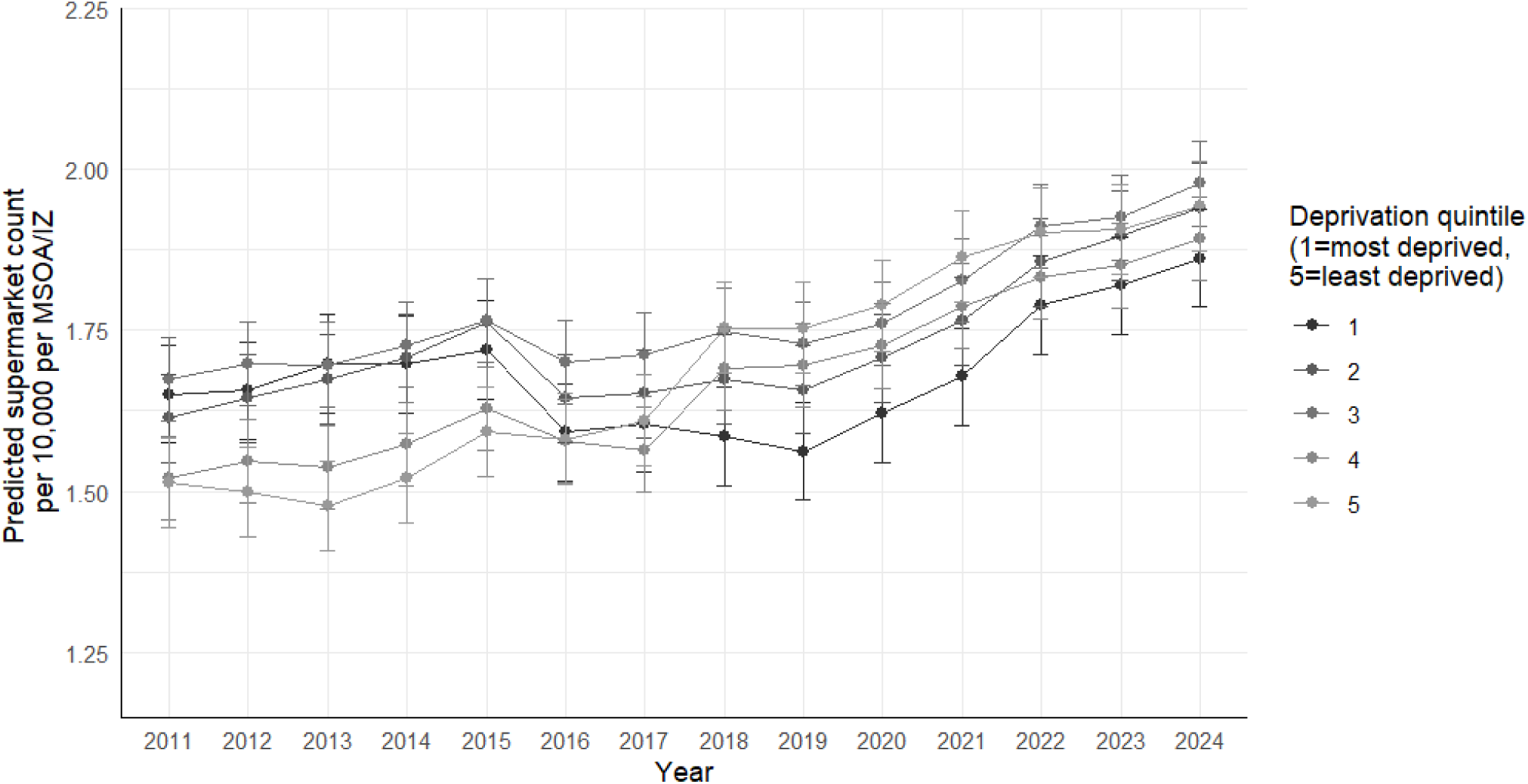
Mean predicted supermarket counts per 10,000 population by deprivation quintile in GB (2011–2024) with 95% CI. Coefficients were derived from linear multilevel models with interaction terms between year and deprivation quintile. Predicted supermarket densities were adjusted for the effect of proportion of population under age 30, proportion of population male and urban/rural classification.

### mRFEI by area-level deprivation

When combining supermarket and fast-food outlet densities in the mRFEI measure, we found that mean mRFEI was consistently lowest in more deprived areas in all years. The 95% CI showed that the most deprived areas (Q1-2) had significantly lower mRFEI than less deprived quintiles (Q3-5) in all years. There were statistically significant differences in mean mRFEI between all quintiles of deprivation (Q1-5) from 2018 onwards. In contrast to the findings for fast-food density, no deprivation quintile showed a statistically significant decrease in mRFEI from 2011 to 2024 (Figure 4).

**Figure 4.**
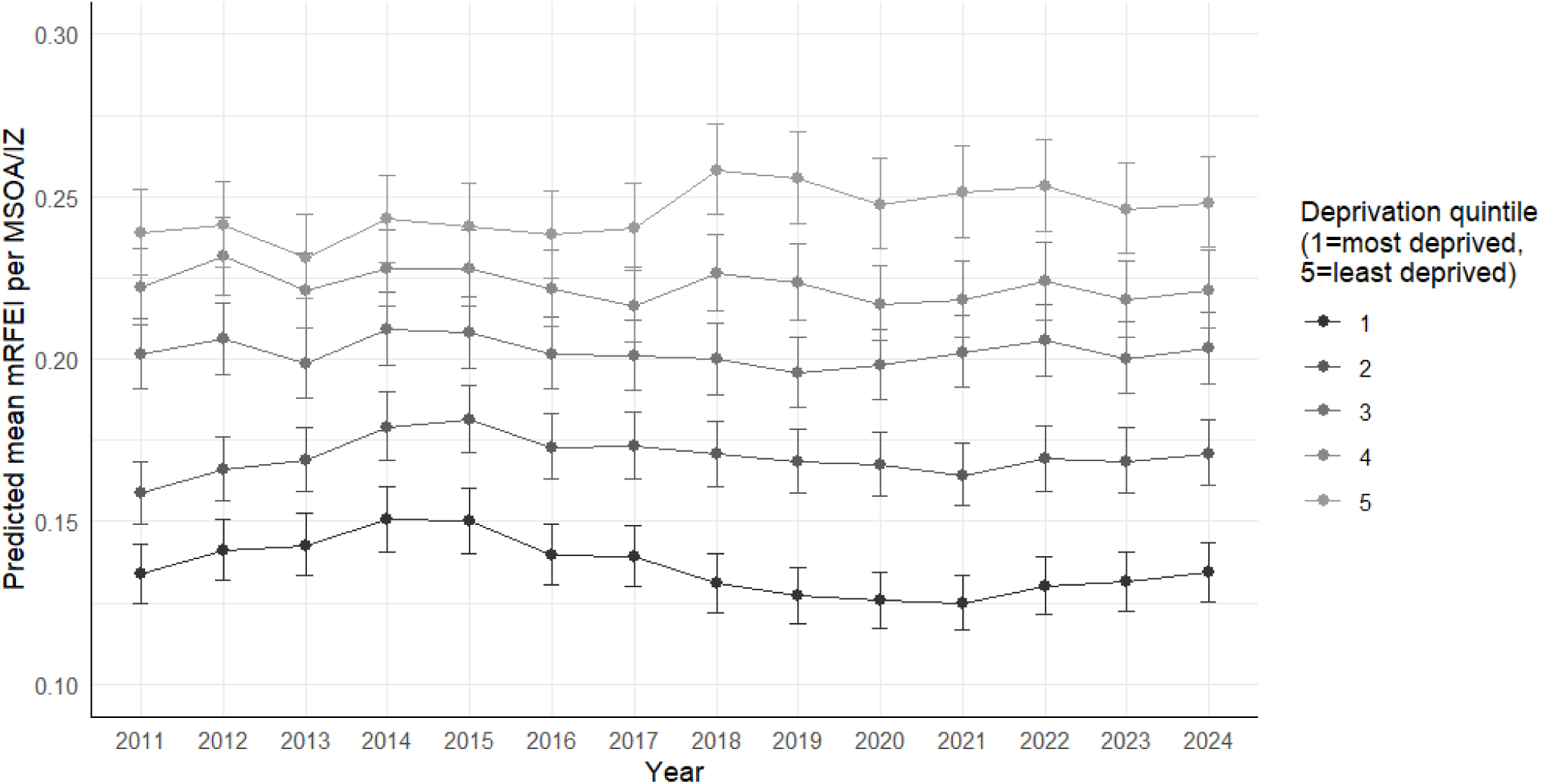
Mean predicted mRFEI per MSOA/IZ by deprivation quintile in GB (2011–2024) with 95% CI. Coefficients were derived from beta-regression models with interaction terms between year and deprivation quintile. Predicted mRFEI are adjusted for the effect of proportion of population under age 30, proportion of population male and urban/rural classification

## Discussion

This novel study was a repeated cross-sectional analysis of area deprivation and food outlet density across GB. We found that the density fast-food outlets increased by 36% and supermarkets increased by 17% across GB from 2011 to 2024. Combining both types of food outlets, we found that the mRFEI decreased from 2011-2024 by 5%, indicative of an increasingly unhealthy RFE overall. Furthermore, we found that more deprived areas consistently had greater fast-food outlet densities compared to less deprived areas, with this disparity growing over the study period by 28%. Although more deprived areas generally had higher densities of supermarkets than less deprived areas, these differences were not statistically significant. When considering fast-food outlets and supermarkets together, more deprived areas consistently had a lower mean mRFEI relative to less deprived areas.

### Food outlet density and deprivation from 2011-2024

Across GB, the average fast-food outlet density was already high in 2011 at 7.0 outlets per 10,000 population, which increased further to 9.5 by 2024.This is equivalent to a 36% increase over 14 years, complementing previous findings by Maguire et al. (2015), who reported a 45% rise in Norfolk over an 18-year period from 1990 to 2008 [8]. Our study also found that more deprived areas had greater densities of fast-food outlets and that this socioeconomic disparity widened over the study period, further echoing previous findings [8]. These findings are also consistent with the literature linking area-level deprivation to higher fast-food outlet density from England and Scotland [8,33–35]. Some repeated cross-sectional studies outside the UK have similarly linked area-level deprivation and the RFE, with similar conclusions regarding the nature of this relationship [36–39].

Our findings indicate that this increase in fast-food outlet density has shown no signs of plateauing or slowing in recent years, suggesting that this trend is likely to continue in future. Furthermore, although deprived areas started with the highest fast-food outlet density in 2011, they experienced the most rapid increases over the subsequent 14 years, underscoring an ongoing intensification of socioeconomically patterned disparities in the food environment. These growing inequalities emphasise the urgency for public health intervention as the continued proliferation of fast-food outlets could exacerbate future obesity prevalence and health inequalities in GB [40].

While supermarkets can offer healthier food options, they are not exclusively providers of nutritious foods, and simply increasing their presence may not be sufficient to improve dietary outcomes. Although we observed a 17% increase in supermarket density over time, this growth did not differ by deprivation level and remained outpaced by the rise in fast-food outlets. As a result, the overall RFE has become increasingly unbalanced, potentially amplifying diet-related health inequalities, particularly in more deprived communities.

This imbalance is further underscored when examining supermarkets and fast-food outlets together using a relative RFE measure, the mRFEI. More deprived areas consistently had lower mRFEI values than less deprived areas, reflecting a comparatively greater prevalence of fast-food outlets relative to supermarkets. This may mean that residents of deprived areas at an inherent disadvantage when attempting to make healthier dietary choices, potentially exacerbating existing socioeconomic disparities in diet quality and obesity [5,41]. While existing research on mRFEI has largely focused on individual-level associations with diet [14,15,42], our study is, to the best of our knowledge, the first in this context to examine longitudinal socioeconomic differences in mRFEI at the area level.

### Policy implications

Fast-food outlets may be targeting deprived areas for expansion because individuals of lower-SEP tend to consume more fast-food [41,43], with underlying causes for this potentially rooted in food insecurity and poor health literacy [5]. In turn, this supply could further encourage fast-food consumption. However, this presents an opportunity to address nationwide obesity rates and socioeconomic disparities in health through interventions targeting the proliferation of fast-food outlets.

Previous UK spatial planning policies have aimed to address public health concerns regarding fast-food takeaway outlets by limiting their proliferation near schools [44], in areas with high outlet density, or where childhood obesity rates are high [45]. However, such policies do not apply nationwide and depend on uptake by individual local authorities, with only about half adopting at least one such policy as of 2019 [46]. Intervention is most needed in deprived areas, therefore policies implementing a proportionate universal approach could produce the most benefit by prioritising intervention in deprived areas [47]. This research supports the need for such policies in line with previous calls for action [3,8,48].

### Strengths and limitations

By conducting a nationwide study, we ameliorate concerns regarding generalisability that may have limited conclusions drawn from previous regional studies in the UK [8] However, since MSOAs in England represent ∼80% of the small areas analysed in this study, trends in Wales and Scotland may have been obscured by those in England.

No differences in supermarket access by area-level deprivation were found, contributing to a mixed evidence base on this relationship. Such inconsistencies may result from differing definitions of a supermarket, for example with some studies combining supermarkets and convenience stores in a single supermarket retail class [12,13] and with some others analysing supermarkets alone [8,37]. Since convenience stores often cluster in deprived areas and primarily sell unhealthy food with limited fresh produce [49], grouping them with supermarkets in this study could have affected our results.

Due to the lack of recent population estimates, 2020/2021 estimates were used for 2021-2024, potentially leading to slight overestimates of food outlet density. However, given the most deprived (Q1) and least deprived (Q5) areas have significantly different fast-food outlet density in all years from 2011, recent fast-food outlet growth would be unlikely to be matched by rates of population growth, and thus likely did not severely affect our conclusions regarding the relationship between area-level deprivation and the RFE.

## Conclusions

Our nationwide study provides key insights into change in the RFE in GB from 2011 to 2024. Notably, the already high fast-food outlet density of 7 outlets per area per 10,000 population on average in 2011, further increased to 9.5 outlets in 2024, equivalent to 36% growth, with no indication of any plateau. In contrast, supermarket density exhibited more modest growth of 17% over the same period. This imbalance between fast-food outlet and supermarket growth was quantified as a 5% decrease in GB’s mean mRFEI score across all small areas over the study period, suggesting that the food environment has become less healthy overall. Lastly, the disparity in fast-food outlet density between more and less deprived areas had widened by 28% over the 14-year period, potentially contributing to an observed exacerbation of diet-related health inequalities over this period. Our findings support the need for further adoption of policies to reduce fast-food outlet density, especially in more deprived communities.

## List of Abbreviations

UK: United Kingdom
GB: Great Britain
RFE: Retail Food Environment
mRFEI: Modified Retail Food Environment Index
OS PoI: Ordnance Survey Points-of-Interest
MSOA: Middle Super Output Area
IZ: Intermediate Zone
LSOA: Lower Super Output Area
DZ: Data Zone
IMD: Index of Multiple Deprivation
CI: Confidence Intervals

## Acknowledgements

We would like to acknowledge those at the Ordnance Survey for providing us with archive OS PoI data on food outlet locations for September 2011, 2012 and 2013.

## Ethics approval

We relied on publicly available data and thus did not require ethical approval.

## Authors’ contributions

AB: Conceptualisation, methodology, formal analysis, data curation, writing original draft, editing; JCH and TB: Conceptualisation, methodology, reviewing and editing, supervision.

## Data availability

Ordnance Survey data are not publicly available, but data access may be permitted upon request. Other data used were in the public domain.

## References

1. Caspi CE, Sorensen G, Subramanian S V., Kawachi I. The local food environment and diet: A systematic review. Health Place. 2012;18:1172–87.

2. Mackenbach JD, Nelissen KGM, Dijkstra SC, Poelman MP, Daams JG, Leijssen JB, et al. A Systematic Review on Socioeconomic Differences in the Association between the Food Environment and Dietary Behaviors. Nutrients 2019, Vol 11, Page 2215 [Internet]. 2019 [cited 2024 Mar 3];11:2215. Available from: https://www.mdpi.com/2072-6643/11/9/2215/htm

3. Townshend T, Lake A. Obesogenic environments: current evidence of the built and food environments. 10.1177/1757913916679860 [Internet]. 2017 [cited 2024 Jan 12];137:38–44. Available from: https://journals.sagepub.com/doi/full/10.1177/1757913916679860?casa_token=UtsfWl_CDT 8AAAAA%3Apz56Z_fv_hidO6PN9yTPDz2pp4_1Evd-D7_utj2RsX9zLpsCA0oeodN2qxR5YDkdQSv_UPrmc_bceA

4. Cobb LK, Appel LJ, Franco M, Jones-Smith JC, Nur A, Anderson CAM. The relationship of the local food environment with obesity: A systematic review of methods, study quality, and results. Obesity. 2015;23:1331–44.

5. Janssen HG, Davies IG, Richardson LD, Stevenson L. Determinants of takeaway and fast food consumption: a narrative review. Nutr Res Rev [Internet]. 2018 [cited 2024 Jan 22];31:16–34. Available from: https://www.cambridge.org/core/journals/nutrition-research-reviews/article/determinants-of-takeaway-and-fast-food-consumption-a-narrative-review/84FCD3376168AF5B70FBC51B4799ECEF

6. MacDonald L, Ellaway A, Ball K, MacIntyre S. Is proximity to a food retail store associated with diet and BMI in Glasgow, Scotland? BMC Public Health [Internet]. 2011 [cited 2024 Apr 4];11:1–9. Available from: https://link.springer.com/articles/10.1186/1471-2458-11-464

7. Macdonald L, Cummins S, Macintyre S. Neighbourhood fast food environment and area deprivation—substitution or concentration? Appetite. 2007;49:251–4.

8. Maguire ER, Burgoine T, Monsivais P. Area deprivation and the food environment over time: A repeated cross-sectional study on takeaway outlet density and supermarket presence in Norfolk, UK, 1990-2008. 2015 [cited 2023 Dec 5]; Available from: http://creativecommons.org/licenses/by/4.0/

9. Burgoine T, Sarkar C, Webster CJ, Monsivais P. Examining the interaction of fast-food outlet exposure and income on diet and obesity: Evidence from 51,361 UK Biobank participants. International Journal of Behavioral Nutrition and Physical Activity [Internet]. 2018 [cited 2023 Dec 5];15:1–12. Available from: https://ijbnpa.biomedcentral.com/articles/10.1186/s12966-018-0699-8

10. Burgoine T, Forouhi NG, Griffin SJ, Wareham NJ, Monsivais P. Associations between exposure to takeaway food outlets, takeaway food consumption, and body weight in Cambridgeshire, UK: population based, cross sectional study. BMJ [Internet]. 2014 [cited 2024 Jan 9];348. Available from: https://www.bmj.com/content/348/bmj.g1464

11. Burgoine T, Forouhi NG, Griffin SJ, Brage S, Wareham NJ, Monsivais P. Does neighborhood fast-food outlet exposure amplify inequalities in diet and obesity? A cross-sectional study. American Journal of Clinical Nutrition [Internet]. 2016 [cited 2024 Jan 25];103:1540–7. Available from: http://ajcn.nutrition.org/article/S0002916523041837/fulltext

12. Smith DM, Cummins S, Taylor M, Dawson J, Marshall D, Sparks L, et al. Neighbourhood food environment and area deprivation: spatial accessibility to grocery stores selling fresh fruit and vegetables in urban and rural settings. Int J Epidemiol [Internet]. 2010 [cited 2024 Apr 9];39:277–84. Available from: 10.1093/ije/dyp221

13. Williamson S, McGregor-Shenton M, Brumble B, Wright B, Pettinger C. Deprivation and healthy food access, cost and availability: a cross-sectional study. Journal of Human Nutrition and Dietetics [Internet]. 2017 [cited 2024 Apr 9];30:791–9. Available from: https://onlinelibrary.wiley.com/doi/full/10.1111/jhn.12489

14. Clary CM, Ramos Y, Shareck M, Kestens Y. Should we use absolute or relative measures when assessing foodscape exposure in relation to fruit and vegetable intake? Evidence from a wide-scale Canadian study. Prev Med (Baltim). 2015;71:83–7.

15. Pinho MGM, Mackenbach JD, Oppert JM, Charreire H, Bárdos H, Rutter H, et al. Exploring absolute and relative measures of exposure to food environments in relation to dietary patterns among European adults. Public Health Nutr [Internet]. 2019 [cited 2024 May 14];22:1037–47. Available from: https://www.cambridge.org/core/journals/public-health-nutrition/article/exploring-absolute-and-relative-measures-of-exposure-to-food-environments-in-relation-to-dietary-patterns-among-european-adults/81BF2A2AB2661D7587F00B29EA17C2CF

16. Thornton LE, Lamb KE, White SR. The use and misuse of ratio and proportion exposure measures in food environment research. International Journal of Behavioral Nutrition and Physical Activity [Internet]. 2020 [cited 2024 May 22];17:1–7. Available from: https://link.springer.com/articles/10.1186/s12966-020-01019-1

17. Ordnance Survey. Points of Interest | Data Products | OS [Internet]. 2024 [cited 2024 Jun 12]. Available from: https://www.ordnancesurvey.co.uk/products/points-of-interest

18. EDINA University of Edinburgh. Digimap [Internet]. Edinburgh, UK; [cited 2024 Jan 16]. p. Home. Available from: https://digimap.edina.ac.uk/

19. Statista. UK: most popular supermarket chains 2023 | Statista [Internet]. 2024 [cited 2024 May 31]. Available from: https://www.statista.com/statistics/1135764/most-popular-supermarkets-in-the-uk/

20. Statista. Chart: The state of the UK supermarket landscape | Statista [Internet]. 2024 [cited 2024 May 31]. Available from: https://www.statista.com/chart/19075/uk-supermarket-market-share/

21. Kantar. Grocery Market Share – Kantar [Internet]. 2024 [cited 2024 May 31]. Available from: https://www.kantarworldpanel.com/en/grocery-market-share/great-britain

22. Statista. Great Britain: Grocery market share 2024 | Statista [Internet]. 2024 [cited 2024 May 31]. Available from: https://www.statista.com/statistics/280208/grocery-market-share-in-the-united-kingdom-uk/

23. Office for National Statistics. Lower layer Super Output Areas (December 2011) Boundaries EW BFE V3| Open Geography Portal [Internet]. 2023 [cited 2024 May 21]. Available from: https://geoportal.statistics.gov.uk/datasets/3011969ff4e84966b2cbc3b642ae32de_0/explore

24. Scottish government spatial data. Data Zone Boundaries 2011 [Internet]. [cited 2024 May 21]. Available from: https://spatialdata.gov.scot/geonetwork/srv/api/records/7d3e8709-98fa-4d71-867c-d5c8293823f2

25. Welsh Government. Welsh Index of Multiple Deprivation [Internet]. StatsWales. 2019 [cited 2024 May 31]. Available from: https://statswales.gov.wales/Catalogue/Community-Safety-and-Social-Inclusion/Welsh-Index-of-Multiple-Deprivation

26. Scottish Government. Scottish Index of Multiple Deprivation [Internet]. statistics.gov.scot. 2020 [cited 2024 May 21]. Available from: https://statistics.gov.scot/resource?uri= http://statistics.gov.scot%2Fdef%2Fconcept%2Ffolders%2Fthemes%2Fscottish-index-of-multiple-deprivation

27. Department for Levelling Up, Housing and Communities and Ministry of Housing C& LG. English indices of deprivation – GOV.UK [Internet]. GOV.UK. 2020 [cited 2024 May 31]. Available from: https://www.gov.uk/government/collections/english-indices-of-deprivation

28. Lombardo M, Aulisa G, Padua E, Annino G, Iellamo F, Pratesi A, et al. Gender differences in taste and foods habits. Nutr Food Sci. 2020;50:229–39.

29. 29. Office for National Statistics. Rural Urban Classification (2011) of LSOAs in EW | Open Geography Portal [Internet]. Open Geography Portal. 2022 [cited 2024 May 31]. Available from: https://geoportal.statistics.gov.uk/datasets/803b5eba7f6f4c998b7d2c5be6729693_0/explore

30. Scottish Government. Population Estimates Detailed (Current Geographic Boundaries) [Internet]. Scottish Government. 2022 [cited 2024 Jan 23]. Available from: https://statistics.gov.scot/data/population-estimates-detailed-current-geographic-boundaries

31. Esri Inc. ArcGIS Pro [Internet. Redlands, CA: Environmental Systems Research Institute; 2021 [cited 2024 Jan 26]. Available from: https://pro.arcgis.com/en/pro-app/index-geonet-allcontent.html

32. R Core Team. R: A language and environment for statistical computing. Vienna, Austria: R Foundation for Statistical Computing; 2023.

33. Cummins SCJ, McKay L, MacIntyre S. McDonald’s Restaurants and Neighborhood Deprivation in Scotland and England. Am J Prev Med. 2005;29:308–10.

34. Macdonald L, Ellaway A, Macintyre S. The food retail environment and area deprivation in Glasgow City, UK. International Journal of Behavioral Nutrition and Physical Activity [Internet]. 2009 [cited 2024 Jul 1];6:1–7. Available from: https://link.springer.com/articles/10.1186/1479-5868-6-52

35. Macdonald L, Olsen JR, Shortt NK, Ellaway A. Do ‘environmental bads’ such as alcohol, fast food, tobacco, and gambling outlets cluster and co-locate in more deprived areas in Glasgow City, Scotland? Health Place. 2018;51:224–31.

36. Hobbs M, Mackenbach JD, Wiki J, Marek L, McLeod GFH, Boden JM. Investigating change in the food environment over 10 years in urban New Zealand: A longitudinal and nationwide geospatial study. Soc Sci Med. 2021;269:113522.

37. Pinho MGM, Mackenbach JD, Den Braver NR, Beulens JJW, Brug J, Lakerveld J. Recent changes in the Dutch foodscape: Socioeconomic and urban-rural differences. International Journal of Behavioral Nutrition and Physical Activity [Internet]. 2020 [cited 2023 Dec 19];17:1–11. Available from: https://ijbnpa.biomedcentral.com/articles/10.1186/s12966-020-00944-5

38. Díez J, Cebrecos A, Rapela A, Borrell LN, Bilal U, Franco M. Socioeconomic Inequalities in the Retail Food Environment around Schools in a Southern European Context. Nutrients 2019, Vol 11, Page 1511 [Internet]. 2019 [cited 2024 May 29];11:1511. Available from: https://www.mdpi.com/2072-6643/11/7/1511/htm

39. Moore L V., Diez Roux A V. Associations of neighborhood characteristics with the location and type of food stores. Am J Public Health [Internet]. 2006 [cited 2024 Jun 26];96:325–31. Available from: https://ajph.aphapublications.org/doi/10.2105/AJPH.2004.058040

40. Swinburn BA, Sacks G, Hall KD, McPherson K, Finegood DT, Moodie ML, et al. The global obesity pandemic: shaped by global drivers and local environments. The Lancet [Internet]. 2011 [cited 2024 Apr 8];378:804–14. Available from: http://www.thelancet.com/article/S0140673611608131/fulltext

41. Barton KL, Wrieden WL, Sherriff A, Armstrong J, Anderson AS. Trends in socio-economic inequalities in the Scottish diet: 2001–2009. Public Health Nutr [Internet]. 2015 [cited 2024 Apr 4];18:2970–80. Available from: https://www.cambridge.org/core/journals/public-health-nutrition/article/trends-in-socioeconomic-inequalities-in-the-scottish-diet-20012009/4FED284DD0663B138DB5A429FD015567

42. Hoenink JC, Lakerveld J, Rutter H, Compernolle S, De Bourdeaudhuij I, Bárdos H, et al. The Moderating Role of Social Neighbourhood Factors in the Association between Features of the Physical Neighbourhood Environment and Weight Status. Obes Facts [Internet]. 2019 [cited 2024 May 21];12:14–24. Available from: 10.1159/000496118

43. Adams J, Goffe L, Brown T, Lake AA, Summerbell C, White M, et al. Frequency and socio-demographic correlates of eating meals out and take-away meals at home: Cross-sectional analysis of the UK national diet and nutrition survey, waves 1-4 (2008-12). International Journal of Behavioral Nutrition and Physical Activity [Internet]. 2015 [cited 2024 Jan 25];12:1–9. Available from: https://ijbnpa.biomedcentral.com/articles/10.1186/s12966-015-0210-8

44. Rahilly J, Williams A, Chang M, Cummins S, Derbyshire D, Hassan S, et al. Changes in the number and outcome of takeaway food outlet planning applications in response to adoption of management zones around schools in England: A time series analysis. Health Place. 2024;87:103237.

45. Brown H, Xiang H, Albani V, Goffe L, Akhter N, Lake A, et al. No new fast-food outlets allowed! Evaluating the effect of planning policy on the local food environment in the North East of England. Soc Sci Med. 2022;306:115126.

46. Keeble M, Burgoine T, White M, Summerbell C, Cummins S, Adams J. How does local government use the planning system to regulate hot food takeaway outlets? A census of current practice in England using document review. Health Place. 2019;57:171–8.

47. Carey G, Crammond B, De Leeuw E. Towards health equity: A framework for the application of proportionate universalism. Int J Equity Health [Internet]. 2015 [cited 2024 Dec 18];14:1–8. Available from: https://equityhealthj.biomedcentral.com/articles/10.1186/s12939-015-0207-6

48. Adams J. Addressing socioeconomic inequalities in obesity: Democratising access to resources for achieving and maintaining a healthy weight. PLoS Med [Internet]. 2020 [cited 2024 Jan 3];17:e1003243. Available from: https://journals.plos.org/plosmedicine/article?id=10.1371/journal.pmed.1003243

49. Harmer G, Jebb SA, Ntani G, Vogel C, Piernas C. Capturing the Healthfulness of the In-store Environments of United Kingdom Supermarket Stores Over 5 Months (January–May 2019). Am J Prev Med. 2021;61:e171–9.

