## Supplementary Material for "The socioeconomic patterning of Great-Britain’s retail food environment: A repeated cross-sectional study of area-level deprivation and food outlet density from 2011-2024"

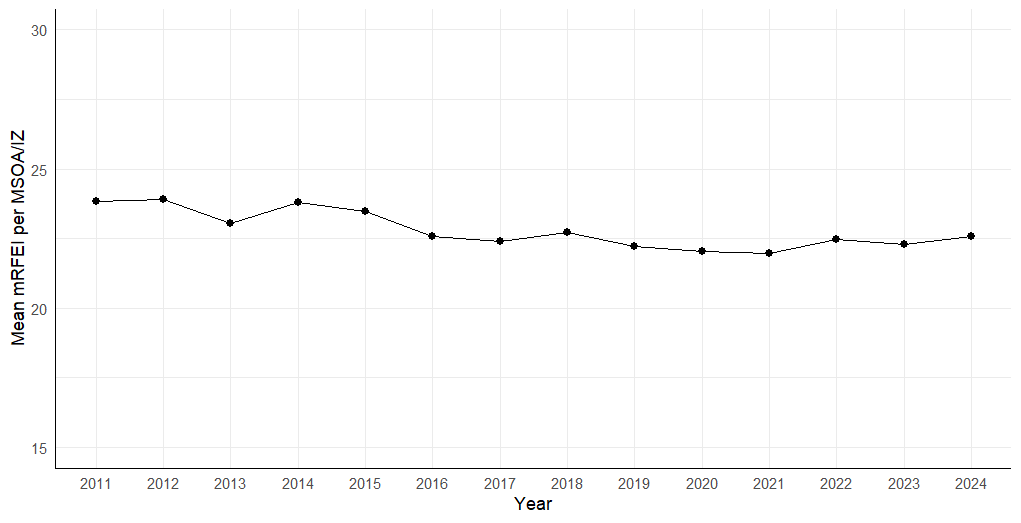

Supplementary Figure 1: Mean mRFEI per MSOA/IZ from 2011-2024 in Great Britain, n=108, 801 observations.

Supplementary Table 1: Mean fast-food outlet and supermarket count per 10,000 population per MSOA/IZ from 2014-2023 by IMD quintile.

| Year | Quintile 1 | | Quintile 2 | | Quintile 3 | | Quintile 4 | | Quintile 5 | |
| --- | --- | --- | --- | --- | --- | --- | --- | --- | --- | --- |
|  | Fast-food outlets | Supermarkets | Fast-food outlets | Supermarkets | Fast-food outlets | Supermarkets | Fast-food outlets | Supermarkets | Fast-food outlets | Supermarkets |
| 2011 | 9.9 | 1.9 | 8.9 | 1.9 | 7.0 | 2.0 | 5.1 | 1.7 | 3.9 | 1.5 |
| 2012 | 10.1 | 1.9 | 9.2 | 2.0 | 7.3 | 2.0 | 5.3 | 1.7 | 4.0 | 1.5 |
| 2013 | 10.6 | 1.9 | 9.6 | 2.0 | 7.6 | 2.0 | 5.5 | 1.7 | 4.2 | 1.5 |
| 2014 | 10.4 | 1.9 | 9.4 | 2.0 | 7.6 | 2.0 | 5.5 | 1.7 | 4.2 | 1.6 |
| 2015 | 10.8 | 1.9 | 9.8 | 2.1 | 7.8 | 2.0 | 5.7 | 1.8 | 4.4 | 1.6 |
| 2016 | 11.0 | 1.8 | 9.9 | 1.9 | 8.1 | 2.0 | 5.9 | 1.7 | 4.5 | 1.6 |
| 2017 | 11.4 | 1.8 | 10.3 | 1.9 | 8.3 | 2.0 | 6.0 | 1.7 | 4.6 | 1.6 |
| 2018 | 12.0 | 1.8 | 10.6 | 2.0 | 8.6 | 2.0 | 6.2 | 1.8 | 4.7 | 1.8 |
| 2019 | 12.4 | 1.7 | 11.0 | 1.9 | 8.7 | 2.0 | 6.4 | 1.8 | 5.0 | 1.8 |
| 2020 | 12.7 | 1.8 | 11.2 | 2.0 | 8.8 | 2.0 | 6.5 | 1.9 | 5.2 | 1.8 |
| 2021 | 13.4 | 1.8 | 11.6 | 2.0 | 9.3 | 2.1 | 6.8 | 1.9 | 5.5 | 1.9 |
| 2022 | 13.3 | 1.9 | 11.8 | 2.1 | 9.3 | 2.2 | 6.9 | 2.0 | 5.6 | 1.9 |
| 2023 | 13.4 | 1.9 | 11.9 | 2.1 | 9.4 | 2.2 | 7.1 | 2.0 | 5.7 | 1.9 |
| 2024 | 13.5 | 2.0 | 11.9 | 2.2 | 9.5 | 2.2 | 7.2 | 2.0 | 5.8 | 2.0 |
